# Search and Retrieval in Dermatology Atlases of Histopathology Images for Risk Stratification of Cutaneous Squamous Cell Carcinoma

**DOI:** 10.64898/2026.01.02.26343356

**Authors:** Ghazal Alabtah, Areej Alsaafin, Saghir Alfasly, Abubakr Shafique, Sobhan Hemati, Anirudh Choudhary, Ravishankar K. Iyer, David DiCaudo, Steve Nelson, Alyssa Stockard, Zach Leibovit-Reiben, Nan Zhang, Krishna R. Kalari, Dennis Murphree, Aaron Mangold, Nneka Comfere, H.R. Tizhoosh

## Abstract

Cutaneous squamous cell carcinoma (cSCC) poses significant clinical challenges due to its rising incidence and potential for metastasis. Histopathologic risk stratification is further limited by substantial inter-observer variability. Unsupervised AI approaches based on content-based image retrieval offer scalable and interpretable decision support for diagnostic pathology.

The objective of this study was to evaluate the use of image retrieval within histopathology atlases to stratify cSCC tumor differentiation from whole-slide images (WSIs), while comparing different patch selection and feature extraction strategies. This retrospective study included 552 archived WSIs comprising 385 well-differentiated, 102 moderately differentiated, and 66 poorly differentiated cases collected across Mayo Clinic sites in Arizona, Florida, and Minnesota.

Image atlases were constructed using multiple patch aggregation strategies (Mosaic, Collage, and Montage) and deep learning encoders (KimiaNet, PathDino, and H-Optimus-0). A leave-one-WSI-out evaluation framework was used to assess differentiation classification performance using accuracy, specificity, sensitivity, and F1 score. Mosaic combined with KimiaNet achieved the highest Top-1 accuracy (74.9%) and specificity (92.6%), while Mosaic with H-Optimus-0 yielded the best Top-5 accuracy (79.0%) and macro-F1 score (62.6%). Collage combined with KimiaNet produced the highest Top-5 specificity (99.5%).

The generalizability of the evaluated AI models varied across hospitals, reflecting differences in imaging protocols, staining practices, and patient populations. Overall, unsupervised image search and retrieval provides effective, annotation-free support for cSCC differentiation and has the potential to enhance dermatopathology workflows when appropriate combinations of patch selection and feature extraction methods are employed.

## 1 Introduction

Cutaneous squamous cell carcinoma (cSCC) is a malignant proliferation of epidermal keratinocytes and most commonly arises in anatomical sites with chronic sun exposure. With increasing incidence rates and a mortality rate comparable to that of melanoma, cSCC is emerging as a significant public health issue [1, 2]. Despite generally favorable clinical outcomes and prognosis, a subset of cSCCs exhibits features associated with an increased risk of recurrence, metastasis, and disease-related death. Accurate identification of tumors with poor prognostic potential is therefore imperative to guide appropriate diagnostic work-up and clinical management.

The Brigham and Women’s Hospital (BWH) classification system focuses on tumor characteristics—including differentiation, diameter, depth, and perineural invasion—to assign risk [3]. However, inter-observer variability in staging remains a major challenge, with an overall upstaging rate of 46.4

Cutaneous squamous cell carcinoma presents a significant clinical challenge due to substantial inter-observer variability in histopathologic risk stratification. Even among experienced dermatopathologists, grading cSCC into well-, moderately-, and poorly differentiated categories is prone to disagreement, reflecting the inherent subjectivity and morphological heterogeneity of these lesions. This variability directly impacts clinical decision-making by influencing treatment intensity, surveillance strategies, and prognostic assessment. Consequently, there is a critical need for more objective and reproducible approaches to cSCC risk stratification.

The recent success of artificial intelligence (AI), particularly deep learning, has opened new horizons in computational pathology [4–7]. Some approaches focus on reducing annotation burden by leveraging eye-tracking data to capture pathologists’ visual attention patterns [8], while others explore few-shot learning strategies for pathology image classification [9].

One of the primary questions that must be addressed before embarking on the design of an AI solution for cSCC—or any other medical application—is the choice between supervised and unsupervised AI approaches [10, 11]. Supervised AI, i.e., models trained for classification and prediction, can often achieve higher levels of accuracy due to extensive training and adaptation to domain-specific subtleties [12]. In contrast, unsupervised AI methods, such as clustering, search, and visualization, do not require task-specific training and may offer improved interpretability [13]. Additionally, supervised AI relies on labeled data (e.g., manual annotations or delineations by pathologists), a requirement that is frequently impractical in medical settings [14].

Beyond traditional supervised and unsupervised paradigms, self-supervised learning has recently attracted significant attention, particularly in the training of large-scale models known as foundation models [15]. Self-Supervised Learning (SSL) leverages unlabeled data to learn general-purpose representations that can be fine-tuned for downstream tasks, thereby reducing the need for extensive manual annotation. However, when applied to computational pathology, self-supervised models often fail to deliver consistently reliable performance. These models tend to exhibit limited diagnostic accuracy, vulnerability to rotation variance, and a strong dependence on extensive domain-specific fine-tuning before they can be used effectively in clinical settings [16–19]. Such limitations underscore the need for pathology-aware pretext tasks and architectures that better capture the structural and morphological invariances inherent to histopathological images.

One of the most promising paradigms within unsupervised AI is search and matching, also referred to as information or image retrieval [20, 21]. Through the <u>indexing</u> of archived cases—comprising previously diagnosed and treated patients—an <u>atlas</u> can be constructed as a structured repository of known cases that encapsulates diagnostic knowledge. New patient cases can then be matched against this atlas to identify morphologically similar cases. Majority voting over the outcomes of the retrieved cases can be used as a form of computational consensus to support decision-making for new patients [22, 23].

The practical implications of employing an atlas for the analysis of squamous cell carcinoma are twofold. First, the atlas can function as a recommendation system for pathologists [23, 24], providing suggested tumor differentiation grades supported by evidence derived from previously diagnosed cases. This evidence-based retrieval not only assists clinical decision-making but also increases diagnostic confidence by anchoring current assessments to historical patterns observed in similar patients. Second, the atlas can serve as a retrieval-augmented generation (RAG) framework to fact-check outputs produced by generative AI models [25, 26]. In this role, the atlas enables <u>source attribution</u>, ensuring that generated narratives or diagnostic summaries can be traced back to verified cases within the archive.

The core objective of this study is to systematically evaluate how different combinations of patch-selection strategies and feature-extraction models influence the construction and performance of whole-slide retrieval maps. Rather than proposing new architectures, our focus is on understanding how these components interact and identifying which pairings yield the most reliable and clinically meaningful retrieval behavior. This comparative analysis provides practical guidance for designing efficient and effective WSI retrieval pipelines.

It is important to emphasize that no new models were trained in this study. Consequently, common concerns related to overfitting or limited generalizability do not apply. Instead, the atlas leverages off-the-shelf models, including foundation models [27], through transfer learning. Embeddings are obtained by freezing the parameters of these pre-existing networks and using their representations directly as feature vectors for search and retrieval. In other words, no new embedding space is learned; rather, the system capitalizes on the representational capacity of existing architectures.

Within this framework, the atlas supports pathologists in grading squamous cell carcinoma by not only recommending a differentiation grade but also justifying the recommendation through retrieved precedent cases [28]. Alternatively, when a pathologist employs a generative AI tool to produce a descriptive or diagnostic report, the atlas can function as a grounding mechanism by attributing the generated output to concrete, retrievable cases. This dual functionality—recommendation and verification—positions the atlas as a practical, transparent, and trustworthy assistant within the dermatopathology workflow.

## 2 Background and History

### 2.1 Building an Atlas

This paper presents experiments evaluating atlases constructed from whole-slide images (WSIs) of cutaneous squamous cell carcinoma (cSCC). Building an atlas requires the extraction of image embeddings (features) from WSI patches, which may contain a mixture of normal tissue, tumor regions, adipose tissue, background, or imaging artifacts. To reduce redundancy and prevent misleading the AI system, an effective patch-selection strategy is essential. Such strategies minimize irrelevant content and direct the model’s attention toward lesion-specific patterns, thereby improving the quality and relevance of the extracted features.

In parallel with patch selection, deep neural networks are employed to extract morphological feature vectors (or <u>embeddings</u>) from each selected WSI patch. These embeddings provide numerical representations of image characteristics, including color, texture, and structural patterns. Tissue similarity can then be assessed by comparing embeddings across patches, where smaller distances correspond to greater morphological similarity. In this study, we evaluate three deep networks for embedding extraction: <u>KimiaNet</u>, a convolutional neural network; <u>PathDino</u>, a vision transformer; and <u>H-Optimus-0</u>, a large foundation model. Other histopathology-trained models could be incorporated within the same framework.

Figure 1 illustrates the key steps involved in atlas construction. The central operation is <u>indexing</u>, in which WSIs are processed and their extracted embeddings are linked to relevant metadata, such as diagnostic reports. When combined with effective patch-selection strategies and robust feature extraction, the resulting atlas can reliably differentiate cSCC cases across varying grades of differentiation. This atlas-based retrieval approach reduces dependence on extensive manual annotation, streamlines analysis, and preserves diagnostic relevance and interpretability.

**Figure 1:**
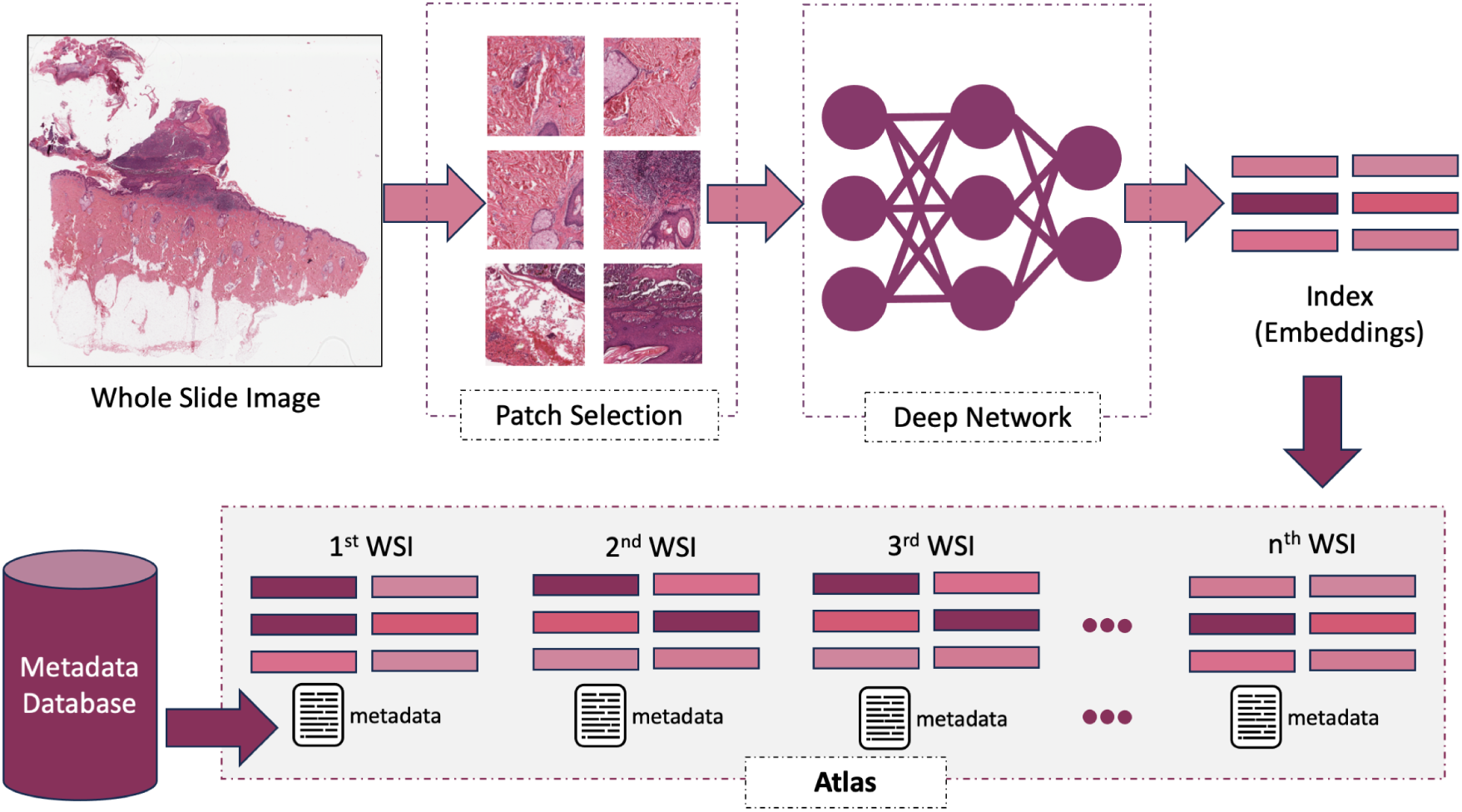
WSI indexing to build an atlas. — Given a WSI, it is decomposed into a small but representative set of patches. These patches are then passed through a trained deep network to extract an embedding (deep feature vector) for each patch. The collection of embeddings associated with a WSI constitutes its WSI index. Relevant metadata (e.g., diagnostic reports) are retrieved from external sources and linked to each WSI index, thereby forming the foundation of the atlas.

An appropriate patch-selection method can highlight image regions that provide critical clues about the degree of differentiation within a WSI. By capturing distinctive color patterns and selecting groups of patches with strong visual uniqueness, such methods guide the model’s attention toward diagnostically relevant tissue regions. A well-trained deep network can then associate these visual patterns with corresponding diagnostic labels, thereby improving the reliability of downstream retrieval and grading.

### 2.2 Patch Selection

Whole-slide image (WSI) patches, also referred to as tiles, are small sub-images that can be processed efficiently by computer algorithms. The principle of <u>Divide Conquer</u> is commonly applied to computationally intensive problems such as gigapixel WSIs: the original large-scale problem (the full WSI) is decomposed into smaller sub-problems (patches), which are analyzed individually and later combined to address the original task [20]. Since processing an entire WSI at once is computationally infeasible, patch-based decomposition is a prerequisite for automated analysis. Effective patching strategies should further aim to select a minimal yet informative subset of patches to enable efficient and scalable search and matching within an atlas.

To ensure broad applicability across different organs, tissue types, and biopsy procedures, patch selection must be unsupervised. Moreover, it should preserve all diagnostically relevant tissue regions without omission [20]. Once a WSI is partitioned into patches, an effective selection strategy is required. The state-of-the-art approach for unsupervised patch selection in image retrieval is the <u>Mosaic</u> method, originally introduced as part of the Yottixel search engine [29]. Yottixel constructs a representative mosaic of patches through a two-stage clustering process followed by a diversity-driven sampling scheme.

In addition to Mosaic, this study investigates two alternative patch-selection methods: the SPLICE framework [30] and the SDM method [31]. SPLICE performs sequential comparisons among WSI patches and retains only non-redundant ones, producing a <u>Collage</u> that captures diverse tissue appearances across the slide. In contrast, <u>Montage</u> selects patches representing distinct morphologies by applying single-centroid *k*-means clustering to WSI patch embeddings extracted using a deep neural network, such as DenseNet [32].

Figure 2 illustrates the general workflow of these three patch-selection methods. Figure 3 provides an example of the SPLICE framework in practice.

**Figure 2:**
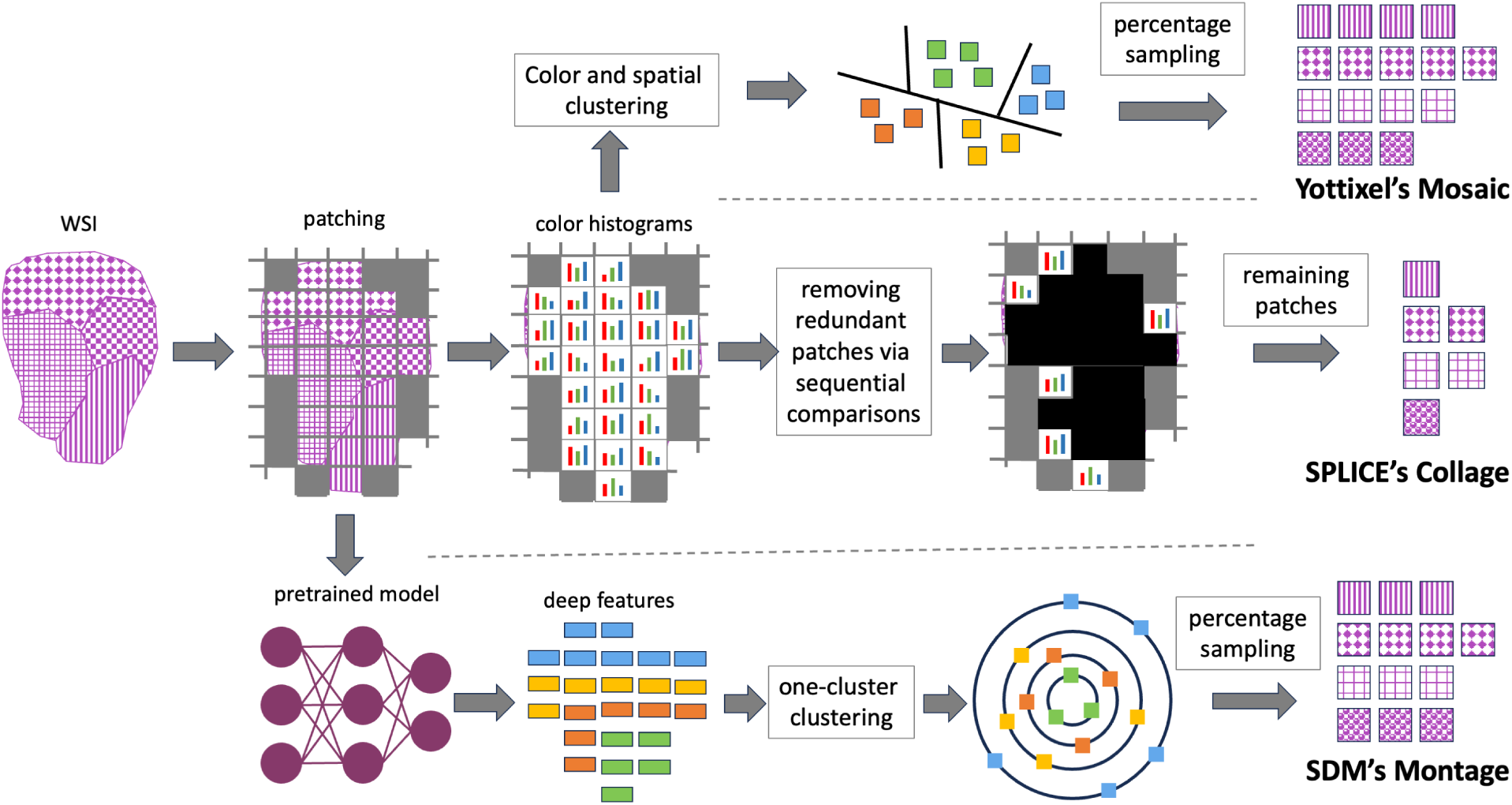
Unsupervised patch selection for WSI indexing and retrieval. Yottixel’s <u>Mosaic</u> (top), SPLICE’s <u>Collage</u> (middle), and SDM’s <u>Montage</u> (bottom) all operate on WSI patches but construct slide-level representations using distinct strategies.

**Figure 3:**
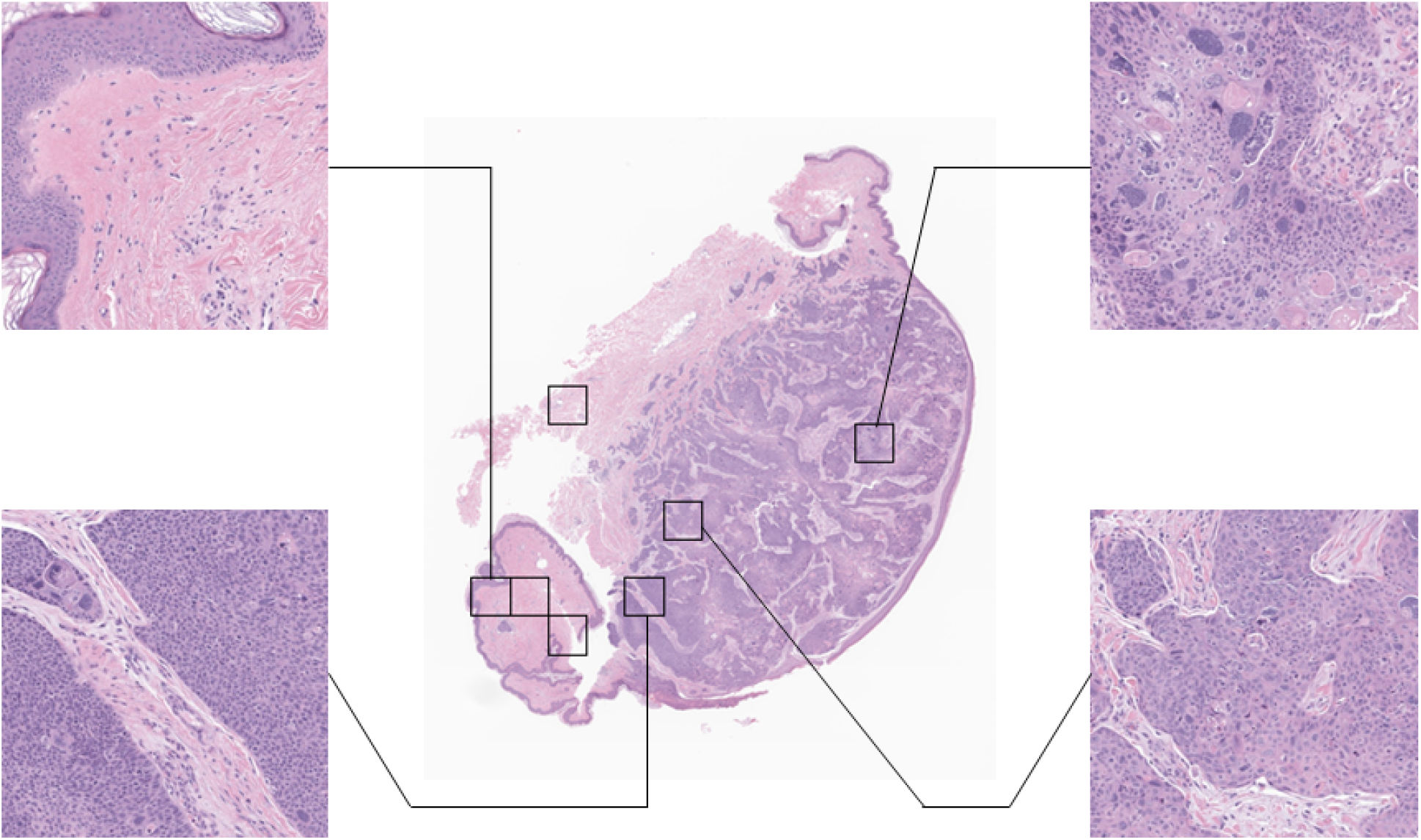
Sample WSI with SPLICE patch selection. Only a subset of patches is magnified for visualization purposes.

All three methods begin by patching the WSI at a lower magnification level, such as 2.5!×, to reduce computational complexity. A segmentation model, such as U-Net [33, 34], is first applied to distinguish tissue from background, retaining only patches that predominantly contain tissue. The U-Net models (with backbones such as MobileNet and EfficientNet-B3) were trained on 244 TCGA whole-slide images spanning multiple organs (e.g., brain, breast, kidney, and lung) [34]. To keep computation tractable, thumbnail images were used instead of full-resolution WSIs. Each thumbnail was associated with a manually refined binary mask indicating tissue versus background, which served as the ground truth for training and evaluating the different U-Net architectures.

Patch selection is then performed exclusively on these tissue-containing regions. Finally, the coordinates of the selected low-magnification patches are mapped to higher magnification levels (e.g., 20× or 40×) to extract the corresponding high-resolution patches for downstream analysis. Patch sizes were selected to match the input requirements of each feature extraction model: PathDINO uses 512 × 512 patches, H-Optimus-0 uses 224 × 224 patches, and KimiaNet uses 1024 × 1024 patches.

Each method includes distinct, customizable parameters that influence both the number and diversity of the selected patches. For Yottixel’s Mosaic, three parameters can be adjusted: (i) the number of clusters used for patch clustering, (ii) the percentage of patches sampled from each cluster, and (iii) the magnification level used for feature extraction. Based on empirical evidence from large-scale studies, these parameters were set to (9, 5, 5), respectively [23, 29]. In SPLICE, patch inclusion is determined by a fixed Euclidean-distance threshold computed from low-magnification (typically 0.625×) color feature vectors. This threshold is an empirically tuned hyperparameter selected to balance redundancy reduction against adequate tissue coverage; we set the SPLICE parameters to (30, 0.062). In contrast, SDM has a single configurable parameter—the feature extraction magnification—which was set to 2.5× to enable fast computation.

### 2.3 Embedding Tissue Patches: Feature Extraction

The next major step in WSI indexing involves using deep networks to extract embeddings from the selected patches, thereby enabling vector-based representations. Specifically, a pre-trained deep network generates embeddings for the selected patches of each WSI. These feature vectors are then used to compute similarities or dissimilarities among WSIs by measuring distances in the embedding space. In essence, this feature extraction stage constitutes the “conquer” phase of our divide conquer pipeline.

We employed three deep networks, each pre-trained specifically for histopathology: <u>KimiaNet</u> [35], a CNN-based DenseNet; <u>PathDino</u> [36], a compact Vision Transformer (ViT); and <u>H-Optimus-0</u> [37], a large Vision Transformer foundation model. The objective of this selection was not to identify a single best-performing model, but rather to include representative architectures from three major categories: convolutional neural networks, Vision Transformers, and large-scale foundation models.

<u>KimiaNet</u> is a fine-tuned DenseNet with approximately 7 million parameters, trained on patches extracted from 7,126 WSIs at 20!× magnification. It emphasizes nuclear distribution and morphology and has demonstrated up to a 44% accuracy improvement across 12 tumor types. For patch-level inference, we followed KimiaNet’s original configuration, using 1024 × 1024 patches at 20× magnification. This magnification applies to feature extraction and may differ from that used during patch selection. Because each patch-selection strategy (Mosaic, Collage, Montage) yields a different number of patches, the resulting WSI-level embedding varies in size. For example, combining KimiaNet with Mosaic, Collage, and Montage produces embedding matrices of size (1024, 88), (1024, 18), and (1024, 99), respectively. KimiaNet outputs a 1,024-dimensional embedding vector for each patch.

The second network, <u>PathDino</u>, is a lightweight foundation-style model composed of five Vision Transformer blocks. It leverages self-attention mechanisms to model relationships among local image regions and is less prone to overfitting—an important advantage in histopathology applications. PathDino operates on patches of size 224×!224 at 20× magnification and produces embeddings with a dimensionality of 384.

The third network, <u>H-Optimus-0</u>, is a large-scale Vision Transformer foundation model trained on more than 500,000 WSIs, encompassing hundreds of millions of patches from diverse tissue types. With 40 transformer blocks, H-Optimus-0 is substantially larger than PathDino and is well suited for modeling complex morphological patterns, particularly in cancer-related tasks. It has achieved an average balanced accuracy of 83% and an average F1 score of 0.822 [38]. Similar to PathDino, H-Optimus-0 accepts 224 × 224 patches at 20× magnification, but generates higher-dimensional embeddings of size 1,536.

Tables 1, 2, 3, and 4 summarize the storage requirements as well as the feature extraction and search times for each combination of patch-selection method and feature extraction model. Detailed descriptions, advantages, limitations, and recommended use cases for the three patch-selection strategies and the three feature extraction models are provided in Table A-1 in the Appendix.

**Table 1:** Storage size (in Megabyte) of atlas indexing for each patching method and feature extraction model.

| Patching Method | KimiaNet | PathDINO | H-Optimus-0 |
| --- | --- | --- | --- |
| SPLICE Collage | 20.9 | 7.9 | 31.3 |
| Yottixel Mosaic | 27.0 | 12.2 | 40.1 |
| SDM Montage | 81.8 | 30.9 | 123.6 |

**Table 2:** Total feature extraction time (in minutes) of the atlas for each model and patch selection method combination.

| Patching Method | KimiaNet | PathDINO | H-Optimus-0 |
| --- | --- | --- | --- |
| SPLICE Collage | 13.43 | 14.41 | 16.30 |
| Yottixel Mosaic | 14.60 | 18.70 | 18.87 |
| SDM Montage | 22.66 | 22.37 | 21.68 |

**Table 3:** Total search time (in minutes) using Leave-One-Out on pre-extracted embeddings from the atlas for each model and feature selection method combination.

| Patching Method | KimiaNet | PathDINO | H-Optimus-0 |
| --- | --- | --- | --- |
| SPLICE Collage | 12.12 | 12.35 | 12.62 |
| Yottixel Mosaic | 17.33 | 17.77 | 17.70 |
| SDM Montage | 84.22 | 86.08 | 86.10 |

**Table 4:** Average search time per WSI in seconds (patch selection + feature extraction + search).

| Patching Method | KimiaNet | PathDINO | H-Optimus-0 |
| --- | --- | --- | --- |
| SPLICE Collage | 3.71 | 21.46 | 15.95 |
| Yottixel Mosaic | 4.26 | 17.10 | 16.71 |
| SDM Montage | 9.13 | 21.46 | 21.26 |

### 2.4 Searching the Atlas

The objective of constructing and using an atlas is to enable fast and efficient search and retrieval of cases that are morphologically similar to a given query image, thereby supporting diagnostic and therapeutic inference for the query patient based on prior evidence. To this end, we compare the morphological characteristics of the query image—quantified through embeddings generated by the three deep models—with those of the images stored in the atlas.

Because each WSI in the atlas is represented by a set of embeddings corresponding to its selected patches, similarity computation is performed at the patch level. Specifically, we compute the Euclidean distances between the embeddings of all query patches and the embeddings associated with every WSI in the atlas. To compare two sets of patches, we adopt a <u>median of minimum distances</u> strategy (see Figure 4).

**Figure 4:**
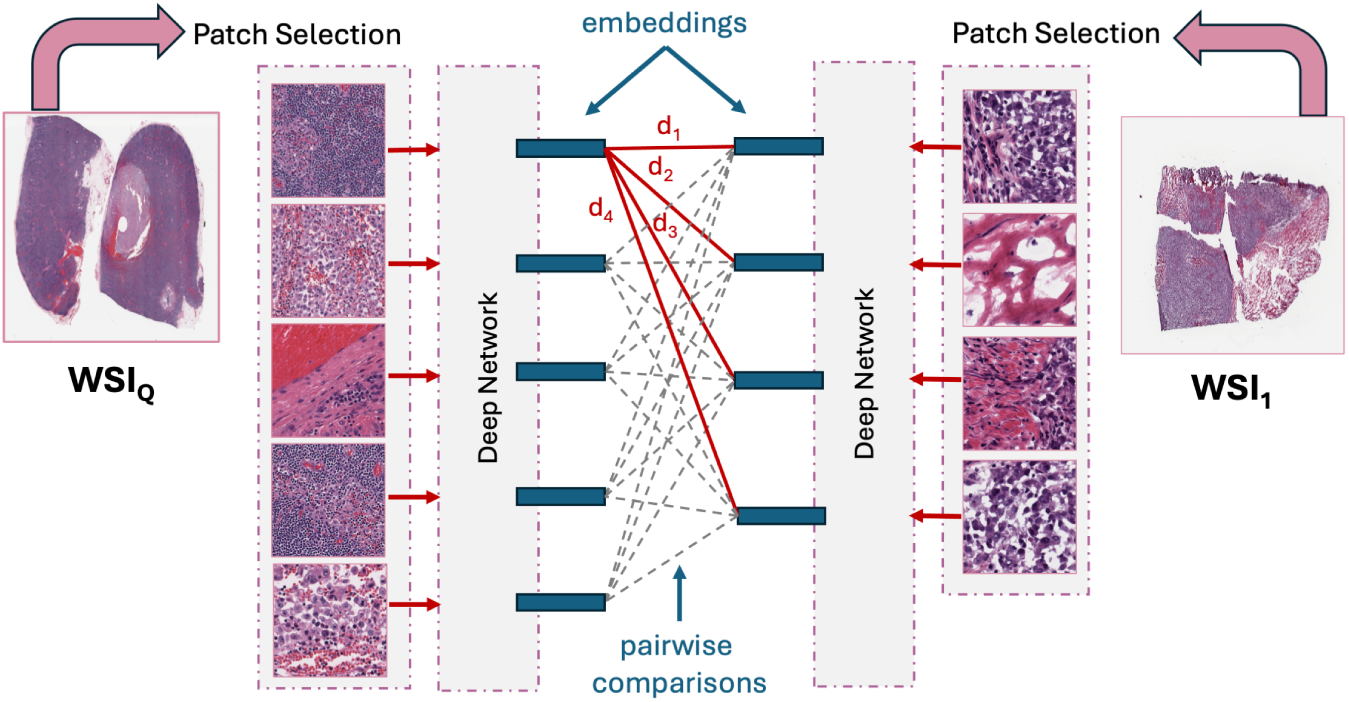
We use the median of minimum distances to measure the similarity between two WSIs. Specifically, the embeddings of the selected patches from a query WSI are compared against the embeddings of patches from a candidate WSI in the atlas. For instance, the first embedding of the query WSI is compared to all embeddings of a candidate WSI, yielding distances *d*_1_ to *d*_4_ (illustrated by the red connections). The minimum value among *d*_1_, *d*_2_, *d*_3_, *d*_4_ is then selected. This process is repeated for all remaining query patches (shown as gray dashed connections), producing a set of minimum distances—one per query patch. The median of these minimum values is used as the overall similarity measure between the two WSIs. [Left image: TCGA-ER-A19M-01Z-00-DX1; right image: TCGA-ER-A19M-06A-06-TSF]

**Figure 5:**
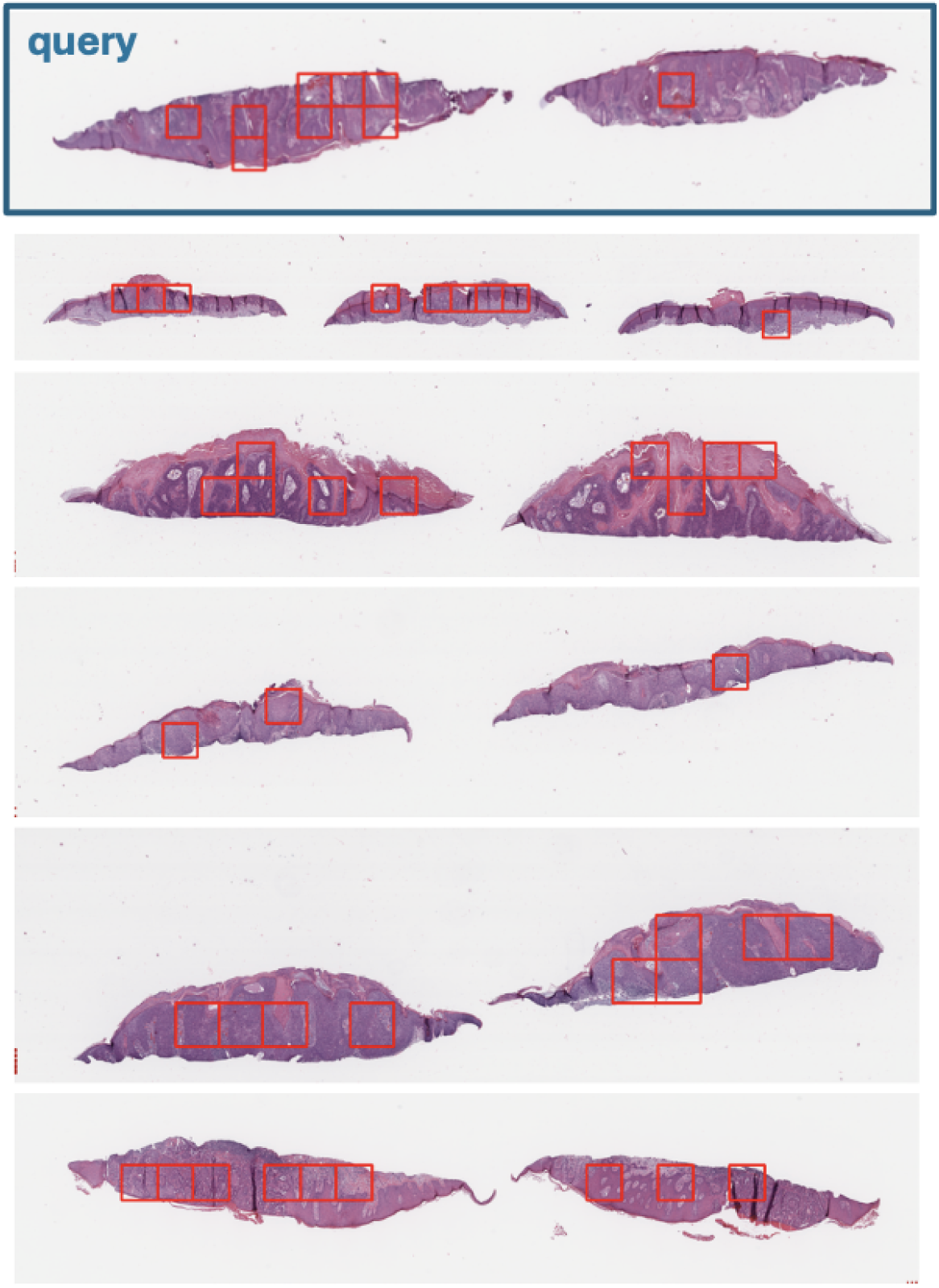
Retrieval example for a misclassified case, showing the selected patches for each retrieved case. Although the query WSI is poorly differentiated, the 1st, 2nd, and 5th retrieved cases are moderately differentiated, resulting in an incorrect majority decision.

**Figure 6:**
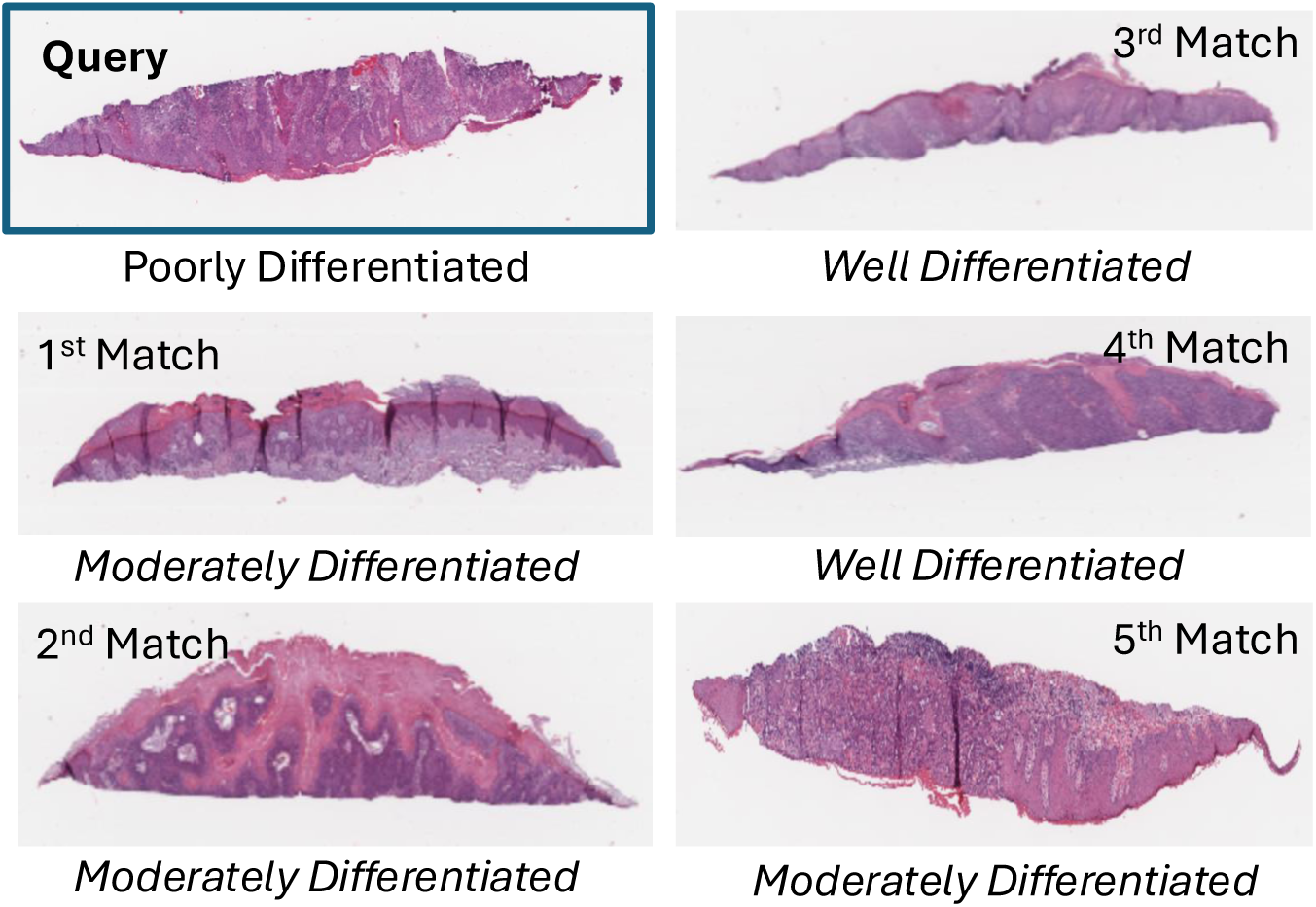
Figure 5 illustrates a misclassified retrieval example (with one representative fragment shown for each WSI to facilitate visual inspection). The query case is poorly differentiated but is misclassified because three of the top five retrieved matches are moderately differentiated. Distinguishing between poorly and moderately differentiated tissue is inherently challenging in dermatopathology, as grading relies on subjec-tive interpretation of histological features and lacks a single standardized grading system. Consequently, substantial inter-observer and intra-observer variability exists among pathologists [40, 41].

Formally, the distance between a query WSI *I_q_* and a candidate WSI *I* is defined as the median of the minimum Euclidean distances between each embedding in the patch set of *I_q_* and the patch set of *I*. In other words, for each query patch, the closest matching patch in *I* is identified, and the median of these minimum distances is used as the overall dissimilarity measure between the two WSIs. The best-matching WSI is therefore the one that minimizes this median distance across all atlas entries.

The embeddings generated by AI models must be evaluated within a reliable and adaptable search framework. Such a search engine should satisfy several key requirements: (1) it must allow seamless integration of different AI models without necessitating significant architectural changes or extensive ablation studies; (2) it must employ an unsupervised patch-selection strategy, thereby eliminating the need for retraining when extending the search to additional organs or tissue types; (3) it should support whole-slide image (WSI) retrieval by leveraging all selected patches without requiring an explicit aggregation step; and (4) it must demonstrate storage efficiency, fast search performance, and high retrieval accuracy compared with alternative solutions.

Based on these criteria, we selected Yottixel as our evaluation platform [20, 29, 39]. Yottixel’s flexible topology enables seamless integration of diverse deep learning models alongside unsupervised patch-selection strategies. This flexibility made it particularly well suited for our retrieval experiments, as it preserved architectural integrity while accommodating new models and patch-selection methods without modification.

## 3 Experiments and Findings

The cSCC cases were identified from two Mayo Clinic retrospective databases comprising curated high-risk cutaneous squamous cell carcinoma cohorts (IRB ID: 21-012833; Full Title: <u>Clinicopathologic and Multi-Omic</u> <u>Stratificati</u>o<u>n of Cutaneous Squamous Cell Carcinoma</u>). Cases were initially retrieved using enterprise-wide internal text-based search engines across Mayo Clinic sites in Arizona, Florida, and Minnesota, selecting specimens with a histopathologic diagnosis of cSCC. A key inclusion criterion was tumor stage T2a or higher according to the Brigham and Women’s Hospital (BWH) staging system.

Following case identification, the corresponding pathology slides were obtained and re-reviewed by board-certified dermatopathologists to confirm the diagnosis of primary cSCC and to verify tumor characteristics, including differentiation, depth, perineural invasion, and extent of tumor invasion. Cases with diagnostic uncertainty were independently reviewed by two dermatopathologists. When discrepancies were identified between the original pathology report and the re-review, the findings from the re-review were considered authoritative and used for subsequent analyses.

Dermatopathology review was also used to select the tissue block that best demonstrated relevant staging features, with particular emphasis on tumor differentiation, while ensuring sufficient tissue and tumor volume for whole-slide imaging. High-throughput slide scanning was performed at 40!× magnification at the Mayo Clinic Rochester Pathology Research Core using Aperio AT Turbo, Aperio AT2, or Aperio GT450 scanners. All digitized slides underwent quality control assessment prior to inclusion in the study.

Matched archival control tissue was obtained from negative surgical margins of excised primary tumors. Dermatopathologic review of these margins confirmed the presence of epidermis without evidence of pathological abnormality. Tissue meeting these criteria was included in the study and labeled as “normal” tissue.

### 3.1 Results

In this section, we evaluate the effectiveness of atlas-based retrieval for cSCC risk stratification. Performance is assessed using standard statistical metrics, including accuracy, sensitivity, specificity, and the macro-averaged F1 score. The F1 score is defined as

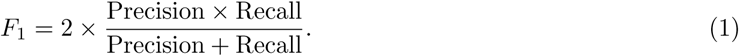

The use of the <u>macro-averaged</u> F1 score is particularly important in this study, as it accounts for class imbalance by weighting each class equally. This consideration is critical for our dataset, in which poorly differentiated cases constitute only approximately 12 A total of 552 cSCC cases were included to evaluate the quality of the experimental atlases and the performance of the search-and-retrieval framework. To better characterize the information captured by each atlas construction method, cases were categorized according to two factors: differentiation level and risk stratification. In this section, we focus on differentiation-based categorization.

Specifically, cases were grouped into three differentiation classes: 1) well-differentiated (385 WSIs), 2) moderately differentiated (102 WSIs), and 3) poorly differentiated (66 WSIs). Tumor differentiation reflects the degree of morphological similarity between malignant cells and the normal squamous cells from which they originate. It is a critical feature in cSCC pathology and staging, as it provides insight into tumor aggressiveness, likelihood of recurrence, and metastatic potential. Pathologists assess differentiation by examining tumor morphology under light microscopy and comparing cellular architecture and cytologic features to those of normal squamous epithelium.

Each case in our dataset was assigned a differentiation label based on dermatopathology review, as defined below:

- **Well-differentiated**: Tumor cells closely resemble normal squamous cells and retain many specialized features. Well-differentiated cSCC typically exhibits slower growth and is associated with a more favorable prognosis.
- **Moderately differentiated**: Tumor cells demonstrate partial loss of differentiation and increased morphological variability compared to normal squamous cells. Moderately differentiated cSCC generally shows more aggressive behavior than well-differentiated tumors.
- **Poorly differentiated**: Tumor cells show minimal resemblance to normal squamous cells and display marked cytologic and architectural atypia. Poorly differentiated cSCC is associated with a higher risk of metastasis and a poorer prognosis.

We constructed nine atlases using all combinations of three patch-selection methods and three deep feature extraction models:

- Mosaic patches with PathDino, KimiaNet, and H-Optimus-0 features
- Collage patches with PathDino, KimiaNet, and H-Optimus-0 features
- Montage patches with PathDino, KimiaNet, and H-Optimus-0 features

Using a <u>leave-one-patient-out</u> validation scheme, we evaluated the performance of all nine configurations for identifying the *top-1* most similar case, as well as for majority voting among the *top-3* and *top-5* retrieved cases. Specifically, top-*k* accuracy is defined as the proportion of test cases for which the correct class label appears within the model’s top *k* highest-ranked predictions. Accordingly, top-1 corresponds to standard accuracy (i.e., the highest-ranked prediction is correct), top-3 indicates that the true label appears among the three most confident predictions, and top-5 extends this criterion to the five most confident predictions.

Table 5 summarizes the evaluation results for all atlas configurations.

- **Top-1 results** — Selecting only the first matched case effectively converts the search-and-retrieval task into a hard classification problem. KimiaNet combined with Mosaic and Collage achieves the highest and second-highest accuracy, respectively. The highest macro-F1 score is obtained by H-Optimus-0 when used with Mosaic patches.
- **Top-3 results** — Considering the top-3 retrieved cases represents a particularly user-friendly scenario, as visual inspection of three similar cases is often both practical and informative in a clinical setting. KimiaNet combined with Montage yields the highest accuracy and specificity, whereas H-Optimus-0 with Mosaic achieves the highest sensitivity and macro-F1 score.
- **Top-5 results** — H-Optimus-0 combined with Mosaic demonstrates the most reliable performance in terms of accuracy, sensitivity, and macro-F1 score. In contrast, KimiaNet combined with Collage produces the highest specificity.

**Table 5:** Comparison of different settings: Best results are highlighted in green, while the second-best results among the nine configurations are highlighted in light blue. Because we employ a leave-one-out validation scheme, predictions from all folds are aggregated into a single test set, yielding one overall performance value per metric.

| Method | Metric | Top-1 | Top-3 | Top-5 |
| --- | --- | --- | --- | --- |
| PathDINO + Montage | Accuracy | 0.721 | 0.768 | 0.777 |
|  | Specificity | 0.925 | 0.966 | 0.989 |
|  | Sensitivity | 0.489 | 0.548 | 0.549 |
|  | F1-macro | 0.504 | 0.560 | 0.544 |
| PathDINO + Mosaic | Accuracy | 0.685 | 0.740 | 0.772 |
|  | Specificity | 0.835 | 0.921 | 0.968 |
|  | Sensitivity | 0.527 | 0.550 | 0.575 |
|  | F1-macro | 0.534 | 0.550 | 0.557 |
| PathDINO + Collage | Accuracy | 0.708 | 0.751 | 0.763 |
|  | Specificity | 0.895 | 0.934 | 0.958 |
|  | Sensitivity | 0.444 | 0.494 | 0.524 |
|  | F1-macro | 0.552 | 0.563 | 0.556 |
| H-Optimus-o + Montage | Accuracy | 0.702 | 0.734 | 0.772 |
|  | Specificity | 0.871 | 0.912 | 0.950 |
|  | Sensitivity | 0.529 | 0.542 | 0.572 |
|  | F1-macro | 0.539 | 0.559 | 0.594 |
| H-Optimus-o + Mosaic | Accuracy | 0.700 | 0.757 | 0.790 |
|  | Specificity | 0.861 | 0.935 | 0.958 |
|  | Sensitivity | 0.700 | 0.757 | 0.790 |
|  | F1-macro | 0.555 | 0.587 | 0.626 |
| H-Optimus-o + Collage | Accuracy | 0.704 | 0.743 | 0.764 |
|  | Specificity | 0.863 | 0.916 | 0.938 |
|  | Sensitivity | 0.543 | 0.562 | 0.564 |
|  | F1-macro | 0.539 | 0.560 | 0.583 |
| KimiaNet + Montage | Accuracy | 0.720 | 0.779 | 0.767 |
|  | Specificity | 0.911 | 0.987 | 0.992 |
|  | Sensitivity | 0.474 | 0.530 | 0.501 |
|  | F1-macro | 0.505 | 0.559 | 0.513 |
| KimiaNet + Mosaic | Accuracy | 0.749 | 0.773 | 0.779 |
|  | Specificity | 0.926 | 0.979 | 0.992 |
|  | Sensitivity | 0.534 | 0.548 | 0.550 |
|  | F1-macro | 0.562 | 0.551 | 0.554 |
| KimiaNet + Collage | Accuracy | 0.730 | 0.758 | 0.764 |
|  | Specificity | 0.934 | 0.984 | 0.995 |
|  | Sensitivity | 0.608 | 0.618 | 0.649 |
|  | F1-macro | 0.503 | 0.502 | 0.501 |

**Analysis of Results** — KimiaNet combined with Mosaic or Collage consistently achieves the highest top-1 accuracy and specificity. This behavior can be attributed to KimiaNet’s DenseNet-based CNN architecture, which produces highly stable, locally textured representations that align well with the abundant low-risk tissue regions typically selected by Mosaic and Collage. In contrast, PathDINO (a lightweight ∼9M-parameter Vision Transformer) and H-Optimus-0 (a substantially larger foundation-model Vision Transformer) rely on global self-attention mechanisms, making their embeddings more sensitive to staining variability and cross-scale inconsistencies, and consequently less stable when modeling dominant, majority-class morphologies. The consistently lower sensitivity observed across all models reflects the pronounced class imbalance in the dataset: high-risk categories (moderately and poorly differentiated tumors) are comparatively rare, morphologically heterogeneous, and spatially sparse within WSIs. As a result, these regions are less likely to be captured within the representative patch subsets selected by Mosaic or Collage. Taken together, architectural differences (CNN versus Vision Transformer), model capacity, and severe class imbalance explain both the substantial gap between specificity and sensitivity and the superior performance of the DenseNet-based KimiaNet on prevalent tissue patterns.

## 4 Discussion and Conclusions

Search and retrieval of histopathology images enables flexible exploration of image atlases based on tissue similarity, facilitating the discovery of morphological patterns beyond predefined diagnostic categories. Unlike classification approaches (i.e., supervised learning), search and retrieval does not require training on labeled datasets and can readily adapt to novel or rare image types without retraining models. Moreover, image retrieval offers greater explainability than classification by returning visually similar cases, allowing users to intuitively understand why a particular result was retrieved. In contrast, classification typically produces a single label without visual context, making the underlying rationale more difficult to interpret. This work aims to make a case for atlas-based search and retrieval as a practical and interpretable paradigm for dermatopathology.

Comparative evaluation of atlas performance across different combinations of patch-selection strategies and AI models provides valuable insight into how differentiation levels and risk stratification in cutaneous squamous cell carcinoma (cSCC) can be captured through unsupervised retrieval. Each patch-selection method and deep network examined in this study embodies distinct design principles, objectives, and trade-offs, leading to observable differences in retrieval performance. Our results demonstrate that the choice of patch-selection strategy and feature-extraction model significantly influences retrieval outcomes. Consequently, identifying an optimal configuration may require empirical evaluation tailored to the specific diagnostic task and dataset.

This study presented a systematic evaluation of atlases constructed from whole-slide images (WSIs) of cSCC. Atlas construction involves extracting image embeddings from patches selected within WSIs. These patches may contain normal tissue, tumor regions, background, or imaging artifacts. As a result, unsupervised patch-selection methods are essential to reduce redundancy, prevent irrelevant content from biasing the retrieval process, and guide the model toward diagnostically meaningful regions. In this work, we investigated three unsupervised patch-selection strategies: Mosaic, Collage, and Montage.

Finally, search and retrieval by indexing query cSCC WSIs against atlas cases leverages the diagnostic knowledge embedded in previously reviewed and annotated patients. New cases can be matched to similar atlas entries to identify morphological correspondences, and majority voting among retrieved cases can be used as a form of computational consensus to support diagnostic inference. Together, these findings highlight the potential of atlas-based, unsupervised search and retrieval as an interpretable, flexible, and annotation-efficient framework for supporting dermatopathology workflows.

## 5 Data Availability

Due to institutional and ethical restrictions related to patient privacy, the whole-slide images (WSIs) used in this study cannot be shared publicly. However, a representative subset of image patches derived from these WSIs may be made available upon reasonable request to Dr. Aaron Mangold, subject to appropriate data use agreements and institutional review.

## Appendix Patching and Models

**Table A-1:** Comparison of patching methods (first Table half) and feature extractors (second Table half), including core design principles, advantages, disadvantages, and ideal application scenarios.

|  | Core Design Principle | Advantages | Disadvantages | Best Use Cases |
| --- | --- | --- | --- | --- |
| **Patch Selection: Mosaic** | Selects representative patches spaced across the WSI using low-magnification diversity | • Strong WSI coverage<br>• Preserves morphological variety<br>• Effective for retrieval and classification | • May include redundant regions<br>• Can miss small abnormalities | • Large-scale retrieval<br>• WSI similarity search<br>• Broad-coverage applications |
| **Patch Selection: SPLICE** | Selects patches maximizing color/texture diversity while suppressing redundancy | • Very compact representation<br>• Highly storage-efficient | • May underrepresents rare morphologies<br>• Patch set may be too small for subtle tasks | • Storage-constrained indexing<br>• Rapid screening<br>• Lightweight workflows |
| **Patch Selection: SDM** | Spatially structured sampling preserving local neighborhood consistency | • Good spatial coherence<br>• Balanced coverage | • Larger patch sets<br>• Potential for redundancy | • Classification tasks<br>• Scenarios requiring spatial regularity |
| **KimiaNet** | ~6M params DenseNet (CNN) trained on TCGA, specializing in local texture modeling | • Highly stable on dominant tissue patterns<br>• Excellent specificity<br>• Robust to noise and stain variation | • Less sensitive to rare or subtle features<br>• Larger patch size than most models | • Majority-class dominated datasets<br>• Retrieval systems<br>• Prioritizing stability and specificity |
| **PathDINO** | ~9M params lightweight ViT with global attention and contrastive pre-training on TCGA | • Efficient and fast<br>• Captures contextual cues<br>• Good generalization for its size<br>• rotationally invariant | • Sensitive to stain/texture variability<br>• Slightly less stable on dominant morphologies | • Resource-limited environments<br>• Multi-center datasets<br>• Medium-scale classification |
| **H-Optimus-0** | ~1.1B params large-scale ViT foundation model pretrained on features extracted from more than 500k histopathology slides | • Extremely rich global representations<br>• Strong cross-domain generalization<br>• Captures complex high-level morphology | • Very high computational cost<br>• Can over-smooth fine textural details<br>• Subsetting may suppress rare high-risk patterns | • Large heterogeneous cohorts<br>• High-performance research pipelines<br>• Workflows with ample computational resources |

## References

[1] John G Muzic, Adam R Schmitt, Adam C Wright, Dema T Alniemi, Adeel S Zubair, Jeannette M Olaza-gasti Lourido, Ivette M Sosa Seda, Amy L Weaver, and Christian L Baum. Incidence and trends of basal cell carcinoma and cutaneous squamous cell carcinoma: a population-based study in olmsted county, minnesota, 2000 to 2010. In Mayo Clinic Proceedings, volume 92, pages 890–898. Elsevier, 2017.

[2] Pritesh S Karia, Jiali Han, and Chrysalyne D Schmults. Cutaneous squamous cell carcinoma: estimated incidence of disease, nodal metastasis, and deaths from disease in the united states, 2012. Journal of the American Academy of Dermatology, 68(6):957–966, 2013.

[3] Pritesh S Karia, Anokhi Jambusaria-Pahlajani, David P Harrington, George F Murphy, Abrar A Qureshi, and Chrysalyne D Schmults. Evaluation of american joint committee on cancer, international union against cancer, and brigham and women’s hospital tumor staging for cutaneous squamous cell carcinoma. Journal of Clinical Oncology, 32(4):327, 2014.

[4] Gabriele Campanella, Matthew G Hanna, Luke Geneslaw, Allen Miraflor, Vitor Werneck Krauss Silva, Klaus J Busam, Edi Brogi, Victor E Reuter, David S Klimstra, and Thomas J Fuchs. Clinical-grade computational pathology using weakly supervised deep learning on whole slide images. Nature medicine, 25(8):1301–1309, 2019.

[5] Miao Cui and David Y Zhang. Artificial intelligence and computational pathology. Laboratory Investigation, 101(4):412–422, 2021.

[6] Taher Dehkharghanian, Youqing Mu, Hamid R Tizhoosh, and Clinton JV Campbell. Applied machine learning in hematopathology. International Journal of Laboratory Hematology, 45:87–94, 2023.

[7] Hamid Reza Tizhoosh and Liron Pantanowitz. Artificial intelligence and digital pathology: challenges and opportunities. Journal of pathology informatics, 9(1):38, 2018.

[8] Tianhang Nan, Song Zheng, Siyuan Qiao, Hao Quan, Xin Gao, Jun Niu, Bin Zheng, Chunfang Guo, Yue Zhang, Xiaoqin Wang, et al. Deep learning quantifies pathologists’ visual patterns for whole slide image diagnosis. Nature Communications, 16(1):5493, 2025.

[9] Hao Quan, Xinjia Li, Dayu Hu, Tianhang Nan, and Xiaoyu Cui. Dual-channel prototype network for few-shot pathology image classification. IEEE Journal of Biomedical and Health Informatics, 28(7): 4132–4144, 2024.

[10] Adil Roohi, Kevin Faust, Ugljesa Djuric, and Phedias Diamandis. Unsupervised machine learning in pathology: the next frontier. Surgical Pathology Clinics, 13(2):349–358, 2020.

[11] Subrata Bhattacharjee, Yeong-Byn Hwang, Kouayep Sonia Carole, Hee-Cheol Kim, Damin Moon, Nam-Hoon Cho, and Heung-Kook Choi. Unsupervised, self-supervised, and supervised learning for histopathological pattern analysis in prostate cancer biopsy. In Proceedings of the Future Technologies Conference, pages 1–17. Springer, 2023.

[12] Hooman H Rashidi, Nam K Tran, Elham Vali Betts, Lydia P Howell, and Ralph Green. Artificial intelligence and machine learning in pathology: the present landscape of supervised methods. Academic pathology, 6:2374289519873088, 2019.

[13] Ewen D McAlpine, Pamela Michelow, and Turgay Celik. The utility of unsupervised machine learning in anatomic pathology. American Journal of Clinical Pathology, 157(1):5–14, 2022.

[14] K Benaggoune, Z Al Masry, C Devalland, S Valmary-degano, N Zerhouni, and LH Mouss. Data labeling impact on deep learning models in digital pathology: A breast cancer case study. In Intelligent Vision in Healthcare, pages 117–129. Springer, 2022.

[15] Rayan Krishnan, Pranav Rajpurkar, and Eric J Topol. Self-supervised learning in medicine and health-care. Nature Biomedical Engineering, 6(12):1346–1352, 2022.

[16] Nita Mulliqi, Anders Blilie, Xiaoyi Ji, Kelvin Szolnoky, Henrik Olsson, Sol Erika Boman, Matteo Titus, Geraldine Martinez Gonzalez, Julia Anna Mielcarz, Masi Valkonen, et al. Foundation models–a panacea for artificial intelligence in pathology? arXiv preprint arXiv:2502.21264, 2025.

[17] Saghir Alfasly, Ghazal Alabtah, Sobhan Hemati, Krishna Rani Kalari, Joaquin J Garcia, and HR Tizhoosh. Validation of histopathology foundation models through whole slide image retrieval. Scientific Reports, 15(1):3990, 2025.

[18] Edwin D de Jong, Eric Marcus, and Jonas Teuwen. Current pathology foundation models are unrobust to medical center differences. arXiv preprint arXiv:2501.18055, 2025.

[19] Matous Elphick, Samra Turajlic, and Guang Yang. Are the latent representations of foundation models for pathology invariant to rotation? arXiv preprint arXiv:2412.11938, 2024.

[20] Hamid R Tizhoosh and Liron Pantanowitz. On image search in histopathology. Journal of Pathology Informatics, page 100375, 2024.

[21] Maral Rasoolijaberi, Morteza Babaei, Abtin Riasatian, Sobhan Hemati, Parsa Ashrafi, Ricardo Gonzalez, and Hamid R Tizhoosh. Multi-magnification image search in digital pathology. IEEE Journal of Biomedical and Health Informatics, 26(9):4611–4622, 2022.

[22] Lei Zheng, Arthur W Wetzel, John Gilbertson, and Michael J Becich. Design and analysis of a content-based pathology image retrieval system. IEEE transactions on information technology in biomedicine, 7(4):249–255, 2003.

[23] Shivam Kalra, Hamid R Tizhoosh, Sultaan Shah, Charles Choi, Savvas Damaskinos, Amir Safarpoor, Sobhan Shafiei, Morteza Babaie, Phedias Diamandis, Clinton JV Campbell, et al. Pan-cancer diagnostic consensus through searching archival histopathology images using artificial intelligence. NPJ digital medicine, 3(1):31, 2020.

[24] Yang Nan, Huichi Zhou, Xiaodan Xing, Giorgos Papanastasiou, Lei Zhu, Zhifan Gao, Alejandro F Frangi, and Guang Yang. Revisiting medical image retrieval via knowledge consolidation. Medical Image Analysis, 102:103553, 2025.

[25] Wenqi Fan, Yujuan Ding, Liangbo Ning, Shijie Wang, Hengyun Li, Dawei Yin, Tat-Seng Chua, and Qing Li. A survey on rag meeting llms: Towards retrieval-augmented large language models. In Proceedings of the 30th ACM SIGKDD conference on knowledge discovery and data mining, pages 6491–6501, 2024.

[26] Karen Ka Yan Ng, Izuki Matsuba, and Peter Chengming Zhang. Rag in health care: a novel framework for improving communication and decision-making by addressing llm limitations. Nejm Ai, 2(1): AIra2400380, 2025.

[27] Mieko Ochi, Daisuke Komura, and Shumpei Ishikawa. Pathology foundation models. JMA journal, 8 (1):121–130, 2025.

[28] Ori Ram, Yoav Levine, Itay Dalmedigos, Dor Muhlgay, Amnon Shashua, Kevin Leyton-Brown, and Yoav Shoham. In-context retrieval-augmented language models. Transactions of the Association for Computational Linguistics, 11:1316–1331, 2023.

[29] Shivam Kalra, Hamid R Tizhoosh, Charles Choi, Sultaan Shah, Phedias Diamandis, Clinton JV Campbell, and Liron Pantanowitz. Yottixel–an image search engine for large archives of histopathology whole slide images. Medical Image Analysis, 65:101757, 2020.

[30] Areej Alsaafin, Peyman Nejat, Abubakr Shafique, Jibran Khan, Saghir Alfasly, Ghazal Alabtah, and Hamid R. Tizhoosh. Sequential patching lattice for image classification and enquiry: Streamlining digital pathology image processing. The American Journal of Pathology, 194(10):1898–1912, 2024. ISSN 0002-9440. doi: 10.1016/j.ajpath.2024.06.007.

[31] Abubakr Shafique, Saghir Alfasly, Areej Alsaafin, Peyman Nejat, Jibran A. Khan, and Hamid R. Tizhoosh. Selection of distinct morphologies to divide & conquer gigapixel pathology images. ArXiv, abs/2311.09902, 2023. URL https://api.semanticscholar.org/CorpusID:265221123.

[32] Gao Huang, Zhuang Liu, and Kilian Q. Weinberger. Densely connected convolutional networks. 2017 IEEE Conference on Computer Vision and Pattern Recognition (CVPR), pages 2261–2269, 2016. URL https://api.semanticscholar.org/CorpusID:9433631.

[33] Olaf Ronneberger, Philipp Fischer, and Thomas Brox. U-net: Convolutional networks for biomedical image segmentation. In Nassir Navab, Joachim Hornegger, William M. Wells, and Alejandro F. Frangi, editors, Medical Image Computing and Computer-Assisted Intervention – MICCAI 2015, Cham, 2015. Springer International Publishing.

[34] Abtin Riasatian, Maral Rasoolijaberi, Morteza Babaei, and Hamid R Tizhoosh. A comparative study of u-net topologies for background removal in histopathology images. In 2020 International Joint Conference on Neural Networks (IJCNN), pages 1–8. IEEE, 2020.

[35] Abtin Riasatian. Kimianet: Training a deep network for histopathology using high-cellularity. Master’s thesis, University of Waterloo, 2020.

[36] Saghir Alfasly, Abubakr Shafique, Peyman Nejat, Jibran Khan, Areej Alsaafin, Ghazal Alabtah, and H. R. Tizhoosh. Rotation-agnostic image representation learning for digital pathology. In Proceedings - 2024 IEEE/CVF Conference on Computer Vision and Pattern Recognition, CVPR 2024, Proceedings of the IEEE Computer Society Conference on Computer Vision and Pattern Recognition, pages 11683–11693. IEEE Computer Society, 2024. doi: 10.1109/CVPR52733.2024.01110. Publisher Copyright: © 2024 IEEE.; 2024 IEEE/CVF Conference on Computer Vision and Pattern Recognition, CVPR 2024; Conference date: 16-06-2024 Through 22-06-2024.

[37] C. Saillard et al. H-optimus-0. https://github.com/bioptimus/, 2024. GitHub repository.

[38] Jack Breen, Katie Allen, Kieran Zucker, Lucy Godson, Nicolas M. Orsi, and Nishant Ravikumar. A comprehensive evaluation of histopathology foundation models for ovarian cancer subtype classification. npj Precision Oncology, 9(1):33, 2025. ISSN 2397-768X. doi: 10.1038/s41698-025-00799-8. URL https://doi.org/10.1038/s41698-025-00799-8.

[39] Isaiah Lahr, Saghir Alfasly, Peyman Nejat, Jibran Khan, Luke Kottom, Vaishnavi Kumbhar, Areej Alsaafin, Abubakr Shafique, Sobhan Hemati, Ghazal Alabtah, et al. Analysis and validation of image search engines in histopathology. IEEE Reviews in Biomedical Engineering, 2024.

[40] Claire Diede, Trent Walker, David R Carr, and Kathryn T Shahwan. Grading differentiation in cutaneous squamous cell carcinoma: a review of the literature. Archives of Dermatological Research, 316 (7):434, 2024.

[41] Jessica M Nash, Kathryn T Shahwan, Catherine Chung, Nadia Abidi, Kaila A Buckley, Yevgeniya Gokun, Xueliang Pan, and David R Carr. Grading differentiation in cutaneous squamous cell carcinoma: Evaluation of interrater and intrarater reliability in mohs surgeons and dermatopathologists. Dermatologic Surgery, pages 10–1097, 2022.

